# Cardiac Inflammation after COVID-19 mRNA Vaccines: A Global Pharmacovigilance Analysis

**DOI:** 10.1101/2021.08.12.21261955

**Authors:** Laurent Chouchana, Alice Blet, Mohammad Al-Khalaf, Tahir S. Kafil, Girish Nair, James Robblee, Milou-Daniel Drici, Marie-Blanche Valnet-Rabier, Joëlle Micallef, Francesco Salvo, Jean-Marc Treluyer, Peter P. Liu

## Abstract

**Background:** To counter the COVID-19 pandemic, mRNA vaccines, namely tozinameran and elasomeran, have been authorized in several countries. These next generation vaccines have shown high efficacy against COVID-19 and demonstrated a favorable safety profile. As widespread vaccinations efforts are taking place, incidents of myocarditis and pericarditis cases following vaccination have been reported. This safety signal has been recently confirmed by the European Medicine Agency and the U.S. Food and Drug Administration.

This study aimed to investigate and analyze this safety signal using a dual pharmacovigilance database analysis.

**Methods:** This is as an observational study of reports of inflammatory heart reactions associated with mRNA COVID-19 vaccines reported in the World Health Organization’s global individual case safety report database (up to June 30^th^ 2021), and in the U.S. Vaccine Adverse Event Reporting System (VAERS, up to May 21^st^ 2021). Cases were described, and disproportionality analyses using reporting odds-ratios (ROR) and their 95% confidence interval (95%CI) were performed to assess relative risk of reporting according to patient sex and age.

**Results:** At a global scale, the inflammatory heart reactions most frequently reported were myocarditis (1241, 55%) and pericarditis (851, 37%), the majority requiring hospitalization (n=796 (64%)). Overall, patients were young (median age 33 [21-54] years). The main age group was 18-29 years old (704, 31%), and mostly males (1555, 68%). Pericarditis onset was delayed compared to myocarditis with a median time to onset of 8 [3-21] vs. 3 [2-6] days, respectively (p=0.001). Regarding myocarditis, an important disproportionate reporting in males (ROR, 9.4 [8.3-10.6]) as well as in adolescents (ROR, 22.3 [19.2-25.9]) and 18-29 years old (ROR, 6.6 [5.9-7.5]) compared to older patients were observed.

**Conclusions:** The inflammatory heart reactions, namely myocarditis and pericarditis, have been reported world-wide shortly following COVID-19 mRNA vaccination. An important disproportionate reporting among adolescents and young adults, particularly in males, was observed especially for myocarditis. Guidelines must take this specific risk into account and to optimize vaccination protocols according to sex and age. While the substantial benefits of COVID-19 vaccination still prevail over risks, clinicians and the public should be aware of these reactions and seek appropriate medical attention.

## Introduction

Myocarditis and pericarditis are inflammatory heart conditions that occur in the setting of excessive host immune response to antigenic stimuli, and have diverse outcomes ranging from mild to severe.^1^ They usually occur after viral infections or other stimuli that induce immune system activation.^2^ Recently, viral myocarditis has been widely reported after SARS-CoV-2 infection and subsequent development of coronavirus disease (COVID-19).^3, 4^ In some instances, these inflammatory reactions have been associated with some drug exposure, such anticancer agents, clozapine or vaccines.^5–7^ A descriptive analysis of drug associated myopericarditis spontaneous reports in the United States (1990–2018) identified that about 0.1% of the cases were related to vaccines.^8^ To date, inflammatory cardiac reactions following vaccination have been mostly reported with live-virus vaccines, such as the smallpox vaccine, ^6, 9, 10^ and inactivated influenza vaccines,^11^ Cases reports of myocarditis/pericarditis secondary to next-generation vaccine platforms, used to prevent COVID-19, have been reported. ^12–17^

Vaccination is critical to contain the SARS-CoV-2 pandemic. Messenger RNA (mRNA) vaccines, which have been produced experimentally during the Zika virus outbreak, represent a new generation of vaccines that can be rapidly responsive to emerging viruses, such as SARS-CoV-2.^18^ Pivotal phase 3 randomized clinical trials were conducted on 71,265 volunteers, including 35,654 who were exposed to the active mRNA vaccines.^19, 20^ Up to July 2021, tozinameran (BNT162b2 mRNA, Pfizer-BioNTech) and elasomeran (mRNA 1273, Moderna) have been distributed in 104 and 49 countries worldwide, respectively, and more than 341 million doses have been administered in the U.S.^21^ While these vaccines appear to have a particularly good safety profile, special attention must be paid to potential adverse effects due to their novelty. Rare but severe adverse drug reactions have been highlighted in clinical trials and thereafter reported after COVID-19 mRNA vaccination, such as anaphylaxis or Bell’s palsy.^22–24^ Recently, a growing signal concerning inflammatory heart reactions, which has not been pointed out during clinical trials, has emerged in several regions of the world, including Israel, Europa and the U.S., with early coverage in the press.^25–27^ A small case series in members of the U.S. Military suggested an incidence below one over 100,000 doses administered.^14^ Preliminary analysis by national authorities led to confirm this safety signal and a probable causal relationship in Europe and in the U.S.^28, 29^

In this study, we aim to document the inflammatory heart reactions reported after COVID-19 mRNA vaccination and further analyze this safety signal using a dual pharmacovigilance database analysis based on the World Health Organization’s (WHO) global Individual Case Safety Report database (VigiBase) and the U.S. Vaccine Adverse Event Reporting System (VAERS). The aims of the study are: 1) to describe these cases and their relevant clinical features, and 2) to perform disproportionality analyses to evaluate a potential association of inflammatory heart reactions reporting after mRNA COVID-19 vaccines with patient’s sex and age.

## Methods

### Study Design

This is a retrospective observational study of spontaneous reports of inflammatory heart reactions associated with mRNA COVID-19 vaccines, collected from VigiBase and the VAERS.

### Data Source

VigiBase (https://www.who-umc.org/vigibase/vigibase/) is the unique WHO global database of individual case safety reports (ICSRs).^30^ This database is maintained, deduplicated and deidentified by Uppsala Monitoring Centre (Uppsala, Sweden). VigiBase is the world’s largest pharmacovigilance database, with over 23 million reports of suspected adverse drug reactions gathered from national pharmacovigilance systems and continuously updated. More than 130 countries, over five continents submit spontaneous safety report in this database. Information concerning the reporter, the patient, the suspected and concomitant drugs, the suspected adverse drug reactions and their seriousness, are reported for each case. Reactions are coded using the hierarchical Medical Dictionary for Regulatory Activities (MedDRA) (https://www.meddra.org/).

VAERS (www.vaers.hhs.gov) is the national pharmacovigilance database to monitor the safety of U.S.-licensed vaccines.^31, 32^ It is run by the Centers for Disease Control and Prevention (CDC) and the U.S. Food and Drug Administration (FDA). VAERS is a spontaneous, passive, reporting system. Health-care professionals, vaccine manufacturers, and consumers (patients, parents, and caregivers) can submit reports of suspected adverse drug reactions to VAERS. They include a short narrative with clinical and biological features on the patient medical history, the reaction and its outcome. This short narrative allows data extraction to deeper describe the reports. U.S. reports from VAERS are transmitted to VigiBase on a regular basis.

### Ethics

This study has obtained ethics approval from the Cochin university Hospital institutional review board (number AAA-2021-08039) in conformity with the French laws and regulations.^33^ All of the data used for analysis were de-identified, and only aggregate data are reported.

### Data Extraction and Analysis

Reports including a COVID-19 mRNA vaccine (tozinameran or elasomeran) as the suspected drug in the adverse reaction have been extracted from both databases. From that, cases of inflammatory heart reactions, i.e. myocarditis, myopericarditis, pericarditis, and pleuropericarditis, were retrieved using the *ad-hoc* high-level terms from MedDRA, “non infectious myocarditis” and “non infectious pericarditis”, coded as reactions. Cases included in this study were registered up to May 21^st^ and June 30^th^ 2021, in VAERS and in VigiBase, respectively.

In VigiBase, for each report of interest, the following data were extracted or assessed: demographic features, continent of reporting, type of reporter, month of reporting, patient sex, patient age, time to reaction onset, seriousness, reaction outcome and type of heart inflammatory reaction Serious cases were defined, according to the WHO, as the occurrence of death, life-threatening adverse event, inpatient hospitalisation or prolongation of an existing hospitalisation, significant disability or requirement of intervention to prevent any of these.

In VAERS, for each report of interest, the following data were extracted or assessed: vaccine type, type of heart inflammatory reaction, patient age, sex, COVID-19 prior to vaccination, reaction occurrence after the 1^st^ or 2^nd^ dose of vaccine, onset of the symptoms, type of symptoms, biological and radiological exams, complications, treatment, and hospitalization. Duplicates in VAERS were removed when identified from the short narrative.

### Disproportionality analysis

Disproportionality analyses, also called case – non-case studies are similar to case – control studies but for purpose of pharmacovigilance studies. These studies are nested in a database of spontaneous reports to assess possible disproportionality in reporting. Disproportionality analysis compares the proportion of a specific adverse drug reaction reported in a specific group (e.g. drug exposure, patient age, patient sex) with the proportion of the same adverse drug reaction for a control group.^34–36^ This statistical approach has shown its value to assess relative risks of adverse drug reaction, that are correlated in most cases with results from meta-analyses of clinical trials.^37^ To assess an association between inflammatory heart reactions after mRNA vaccines and patient age or sex, we performed a disproportionality analysis among different patient groups. Characteristics of interest are patient sex and age groups, specially 12-17 years, 18-29 years and over 30 years, as reported by safety signals of concern. Cases are reports concerning a specific patient group and that include the terms myocarditis, pericarditis or pleuropericarditis as reactions after a COVID-19 mRNA vaccine. Non-cases are reports concerning a specific patient group and that include all other reactions to a COVID-19 mRNA vaccine. Therefore, our analyses assessed a potential difference of reporting of inflammatory heart reactions secondary to COVID-19 mRNA vaccines in male compared to female and in 12-17 years or 18-29 years old patients compared to patients over 30 years old. Thereafter, sensitivity analyses were performed, restricted to serious reports only, and to reports originating from a healthcare professional.

### Statistical analysis

Descriptive analysis was performed on the cases retrieved from the two databases. Quantitative variables were expressed as median ± interquartile range (IQR) and compared using non-parametric analysis. Qualitative variables were expressed in number and percentages. Time to reaction onset was analyzed using a survival Mantel-Cox model.

Disproportionality in reporting between groups is expressed using reporting odds ratio (ROR) and its 95% confidence interval (95% CI). ROR is a ratio similar in concept to the odds ratio in case – control studies and corresponds to the exposure odds among reported cases of interest over the exposure odds among reported non-cases. ROR (95% CI) were calculated as 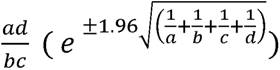, where *a* is the number of cases reported with COVID-19 mRNA vaccines in a specific patient group, *b* is the number of non-cases (i.e., all other adverse drug reactions) reported with COVID-19 mRNA vaccines in a specific patient group, *c* is the number of cases reported with COVID-19 mRNA vaccines in a control group and *d* is the number of non-cases reported with COVID-19 mRNA vaccines in a control group.

## Results

### Demographics and characteristics of cases from the worldwide VigiBase

As of the end of June 2021, of a total of 26,258,646 reports registered in VigiBase, 716,576 reports were related to a COVID-19 mRNA vaccine as a suspected drug, including 495,493 with tozinameran and 221,368 with elasomeran (including 285 with both vaccines). Among them, 2277 cases of inflammatory heart reactions (31.8 per 10,000 of COVID-19 mRNA vaccine reports) were retrieved in the worldwide VigiBase. Cases were largely reported from North America (63.4%) and Europa (35.0%). Of the cases, 1655 (72.7%) were reported with tozinameran and 622 (27.3%) with elasomeran (**Table 1**). The inflammatory heart reactions most commonly reported were myocarditis (1241, 54.5%) and pericarditis (851, 37.4%). Patients were of the younger age bracket (median age 33 [21-54]) mostly from the group between 18-29 year-old (704, 30.9%), more commonly males (1555 (68.3%). Patient distribution was different according to age and sex (p<0.0001) for both myocarditis and pericarditis (**Figure 1**). Specifically, most of myocarditis cases have been reported in 12-17 and 18-29 years old males. Myocarditis cases originating from the U.S. were mainly reported after May 2021, although reactions occur alongside with vaccination achievements (**Figure 2A** and **suppl. figure 1**). Overall, the median time to onset was 3 [2-14] days after vaccine injection. Pericarditis onset was delayed compared to myocarditis with a median time to onset of 8 [3-21] *vs*. 3 [2-6] days, respectively (p=0.001, **Figure 3A**). Drug other than COVID-19 mRNA vaccines were suspected in the cardiac reaction onset in 200 (8.7%) patients. Overall, most of the myocarditis and pericarditis reported cases required hospitalization (1015 (81.8%) and 492 (57.8%), respectively), and 219 (21.5%) and 101 (20.5%) were life threatening, respectively. In 22 patients, including 15 (1.2%) myocarditis cases (median age 60 [56-78] years old), 5 pericarditis cases (median age 71 [67-77] years old) and 2 myopericarditis cases (age 55 and 83), the cardiac inflammatory reaction had a fatal outcome. Among these fatal cases, the vaccine was the only suspected drug in all except one case (an anti-PDL1, atezolizumab, as other suspected drug).

**Figure 1.**
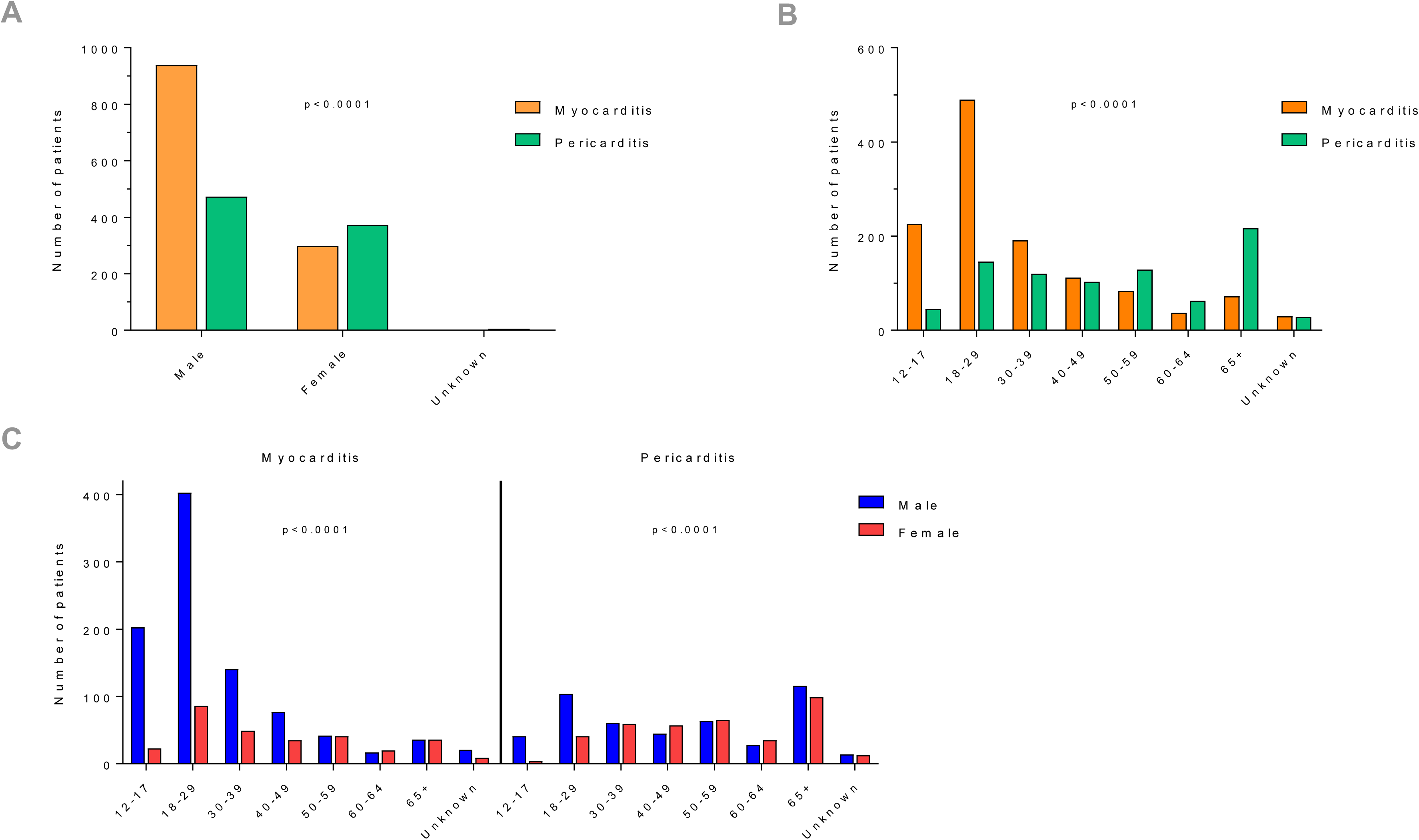
Clinical and demographic features concerning inflammatory heart reaction after mRNA COVID-19 vaccines. The data shown refer to VigiBase data. **(A)** Sex distribution according to reaction type (myocarditis and pericarditis). **(B)** Reaction type (Myocarditis or Pericarditis) distribution according to age. Myocarditis presented most frequently in young people of 18-29 years old, whereas pericarditis presented most frequently in people over 65 years old. **(C)** Sex distribution according to reaction type and age.

**Figure 2.**
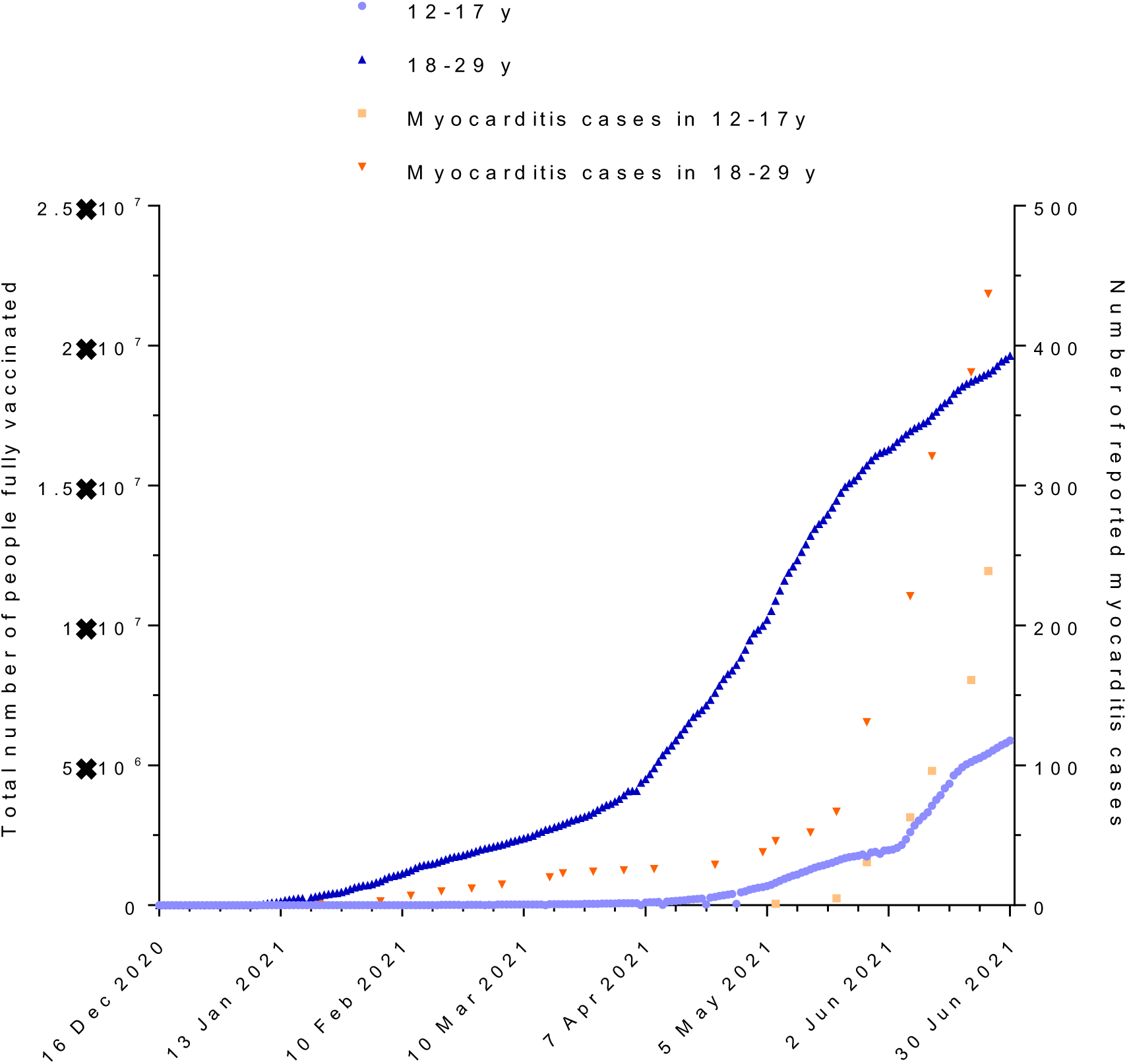
Reported cases of myocarditis and number of people vaccinated among 12-17 and 18-29 year old in the U.S.. Myocarditis originating from the U.S. were mainly reported after May 2021.

**Figure 3.**
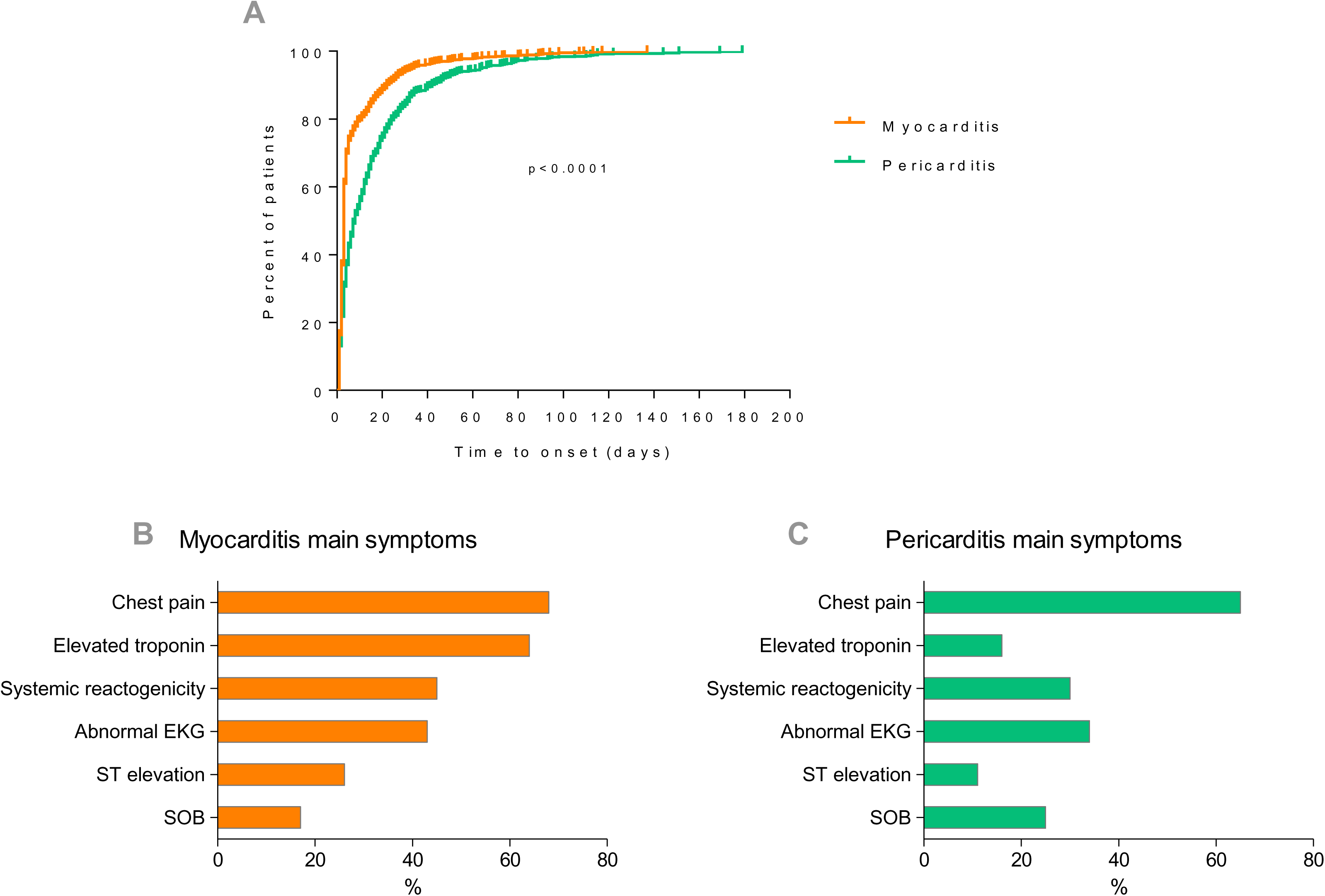
Time to onset and main symptoms reported by type of reaction. Time to onset was earlier for myocarditis. Most frequent symptoms reported were chest pain, elevated troponin and systemic reactogenicity. SOB, shortness of breath

**Table 1.**
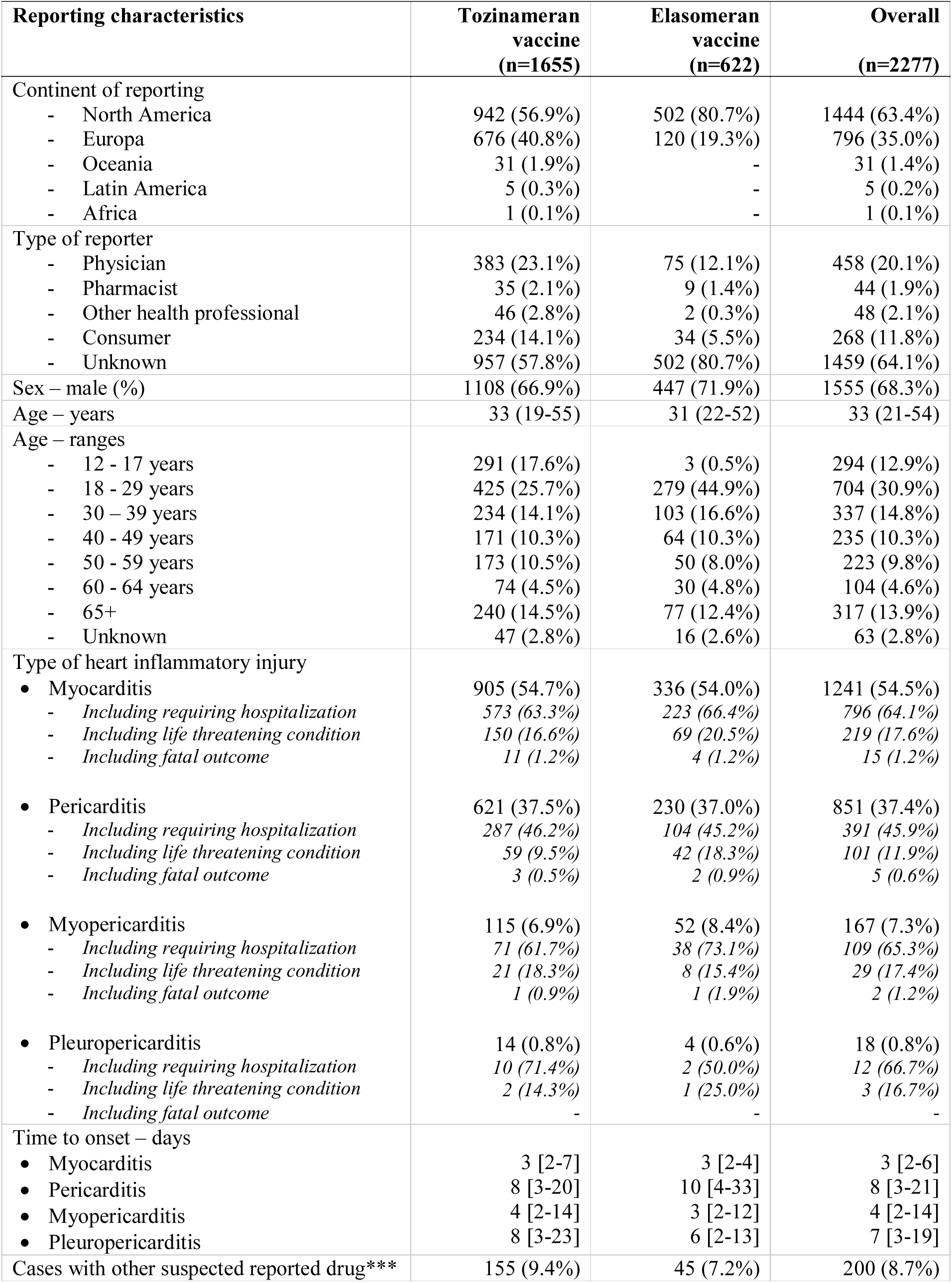
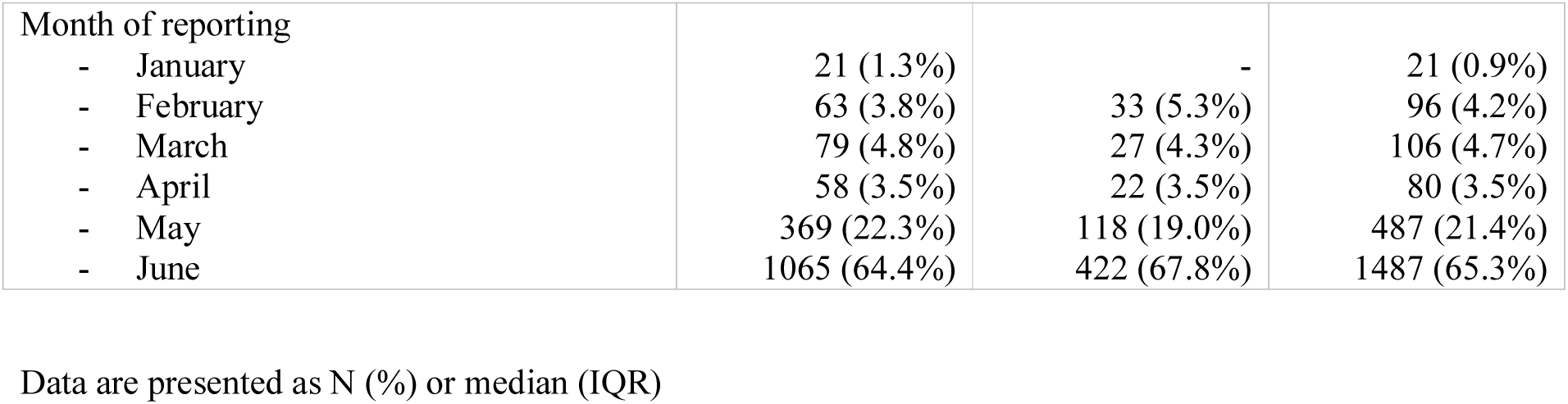
Characteristics of acute inflammatory heart cases reported with COVID-19 mRNA vaccines within the WHO global safety database.

| Reporting characteristics | Tozinameran vaccine<br>(n=1655) | Elasomeran vaccine<br>(n=622) | Overall<br>(n=2277) |
| --- | --- | --- | --- |
| Continent of reporting |  |  |  |
| - North America | 942 (56.9%) | 502 (80.7%) | 1444 (63.4%) |
| - Europa | 676 (40.8%) | 120 (19.3%) | 796 (35.0%) |
| - Oceania | 31 (1.9%) | - | 31 (1.4%) |
| - Latin America | 5 (0.3%) | - | 5 (0.2%) |
| - Africa | 1 (0.1%) | - | 1 (0.1%) |
| Type of reporter |  |  |  |
| - Physician | 383 (23.1%) | 75 (12.1%) | 458 (20.1%) |
| - Pharmacist | 35 (2.1%) | 9 (1.4%) | 44 (1.9%) |
| - Other health professional | 46 (2.8%) | 2 (0.3%) | 48 (2.1%) |
| - Consumer | 234 (14.1%) | 34 (5.5%) | 268 (11.8%) |
| - Unknown | 957 (57.8%) | 502 (80.7%) | 1459 (64.1%) |
| Sex – male (%) | 1108 (66.9%) | 447 (71.9%) | 1555 (68.3%) |
| Age – years | 33 (19-55) | 31 (22-52) | 33 (21-54) |
| Age – ranges |  |  |  |
| - 12 - 17 years | 291 (17.6%) | 3 (0.5%) | 294 (12.9%) |
| - 18 - 29 years | 425 (25.7%) | 279 (44.9%) | 704 (30.9%) |
| - 30 – 39 years | 234 (14.1%) | 103 (16.6%) | 337 (14.8%) |
| - 40 - 49 years | 171 (10.3%) | 64 (10.3%) | 235 (10.3%) |
| - 50 - 59 years | 173 (10.5%) | 50 (8.0%) | 223 (9.8%) |
| - 60 - 64 years | 74 (4.5%) | 30 (4.8%) | 104 (4.6%) |
| - 65+ | 240 (14.5%) | 77 (12.4%) | 317 (13.9%) |
| - Unknown | 47 (2.8%) | 16 (2.6%) | 63 (2.8%) |
| Type of heart inflammatory injury |  |  |  |
| • Myocarditis | 905 (54.7%) | 336 (54.0%) | 1241 (54.5%) |
| - Including requiring hospitalization | 573 (63.3%) | 223 (66.4%) | 796 (64.1%) |
| - Including life threatening condition | 150 (16.6%) | 69 (20.5%) | 219 (17.6%) |
| - Including fatal outcome | 11 (1.2%) | 4 (1.2%) | 15 (1.2%) |
| • Pericarditis | 621 (37.5%) | 230 (37.0%) | 851 (37.4%) |
| - Including requiring hospitalization | 287 (46.2%) | 104 (45.2%) | 391 (45.9%) |
| - Including life threatening condition | 59 (9.5%) | 42 (18.3%) | 101 (11.9%) |
| - Including fatal outcome | 3 (0.5%) | 2 (0.9%) | 5 (0.6%) |
| • Myopericarditis | 115 (6.9%) | 52 (8.4%) | 167 (7.3%) |
| - Including requiring hospitalization | 71 (61.7%) | 38 (73.1%) | 109 (65.3%) |
| - Including life threatening condition | 21 (18.3%) | 8 (15.4%) | 29 (17.4%) |
| - Including fatal outcome | 1 (0.9%) | 1 (1.9%) | 2 (1.2%) |
| • Pleuropericarditis | 14 (0.8%) | 4 (0.6%) | 18 (0.8%) |
| - Including requiring hospitalization | 10 (71.4%) | 2 (50.0%) | 12 (66.7%) |
| - Including life threatening condition | 2 (14.3%) | 1 (25.0%) | 3 (16.7%) |
| - Including fatal outcome | - | - | - |
| Time to onset – days |  |  |  |
| • Myocarditis | 3 [2-7] | 3 [2-4] | 3 [2-6] |
| • Pericarditis | 8 [3-20] | 10 [4-33] | 8 [3-21] |
| • Myopericarditis | 4 [2-14] | 3 [2-12] | 4 [2-14] |
| • Pleuropericarditis | 8 [3-23] | 6 [2-13] | 7 [3-19] |
| Cases with other suspected reported drug*** | 155 (9.4%) | 45 (7.2%) | 200 (8.7%) |
| Month of reporting |  |  |  |
| - January | 21 (1.3%) | - | 21 (0.9%) |
| - February | 63 (3.8%) | 33 (5.3%) | 96 (4.2%) |
| - March | 79 (4.8%) | 27 (4.3%) | 106 (4.7%) |
| - April | 58 (3.5%) | 22 (3.5%) | 80 (3.5%) |
| - May | 369 (22.3%) | 118 (19.0%) | 487 (21.4%) |
| - June | 1065 (64.4%) | 422 (67.8%) | 1487 (65.3%) |

Data are presented as N (%) or median (IQR)

### Disproportionality analysis

Compared to females, vaccination by COVID-19 mRNA vaccines in males was associated with an increased risk of myocarditis and pericarditis reporting with an ROR (95% CI) of 9.4 (8.3-10.6) and 3.7 (3.2-4.2), respectively (**Figure 4A and B**). Analysis according to age group showed a marked disproportionate reporting of myocarditis after COVID-19 mRNA vaccines in 12-17 years old (ROR 22.3 [19.2-25.9]) and 18-29 years old (ROR 6.6 [5.9-7.5]) recipients compared with recipients over 30 years old. Specifically, this increased reporting was more marked in males than in females, among all age groups (**Figure 4A**). Regarding pericarditis, a disproportionate reporting in males among all age groups, compared to females was observed (**Figure 4B**). Younger males belonging to 12-17 and 18-29 years old groups were also more prone to pericarditis reporting after COVID-19 mRNA vaccines compared to over 30 years old group. This finding not being true in females in whom no disproportionality in pericarditis reporting was observed according to age. Further sensitivity analyses restricted to serious reports and to reports originating form healthcare professionals identified consistent results (**Suppl tables S2, S3, S4 and S5**).

**Figure 4.**
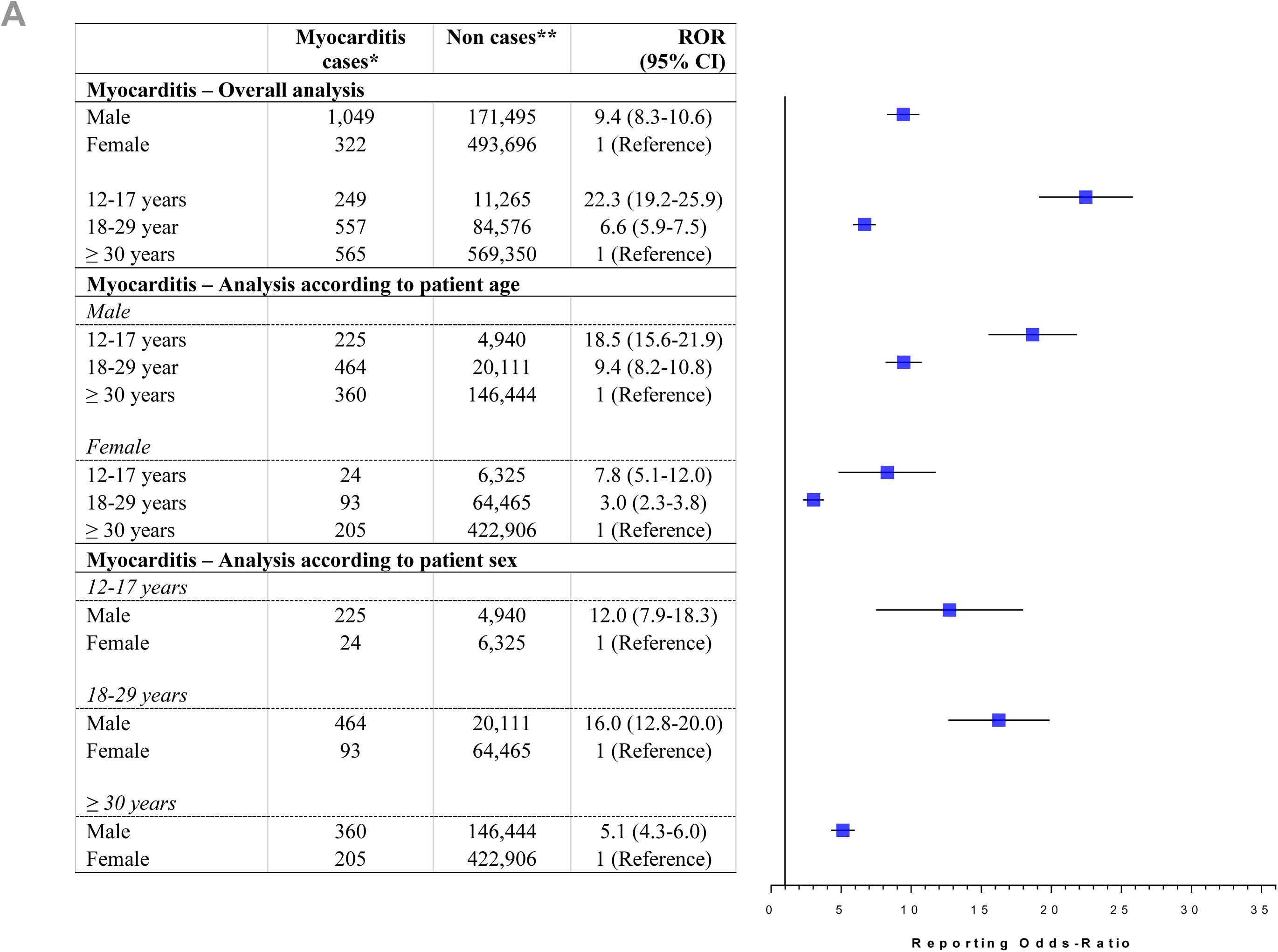
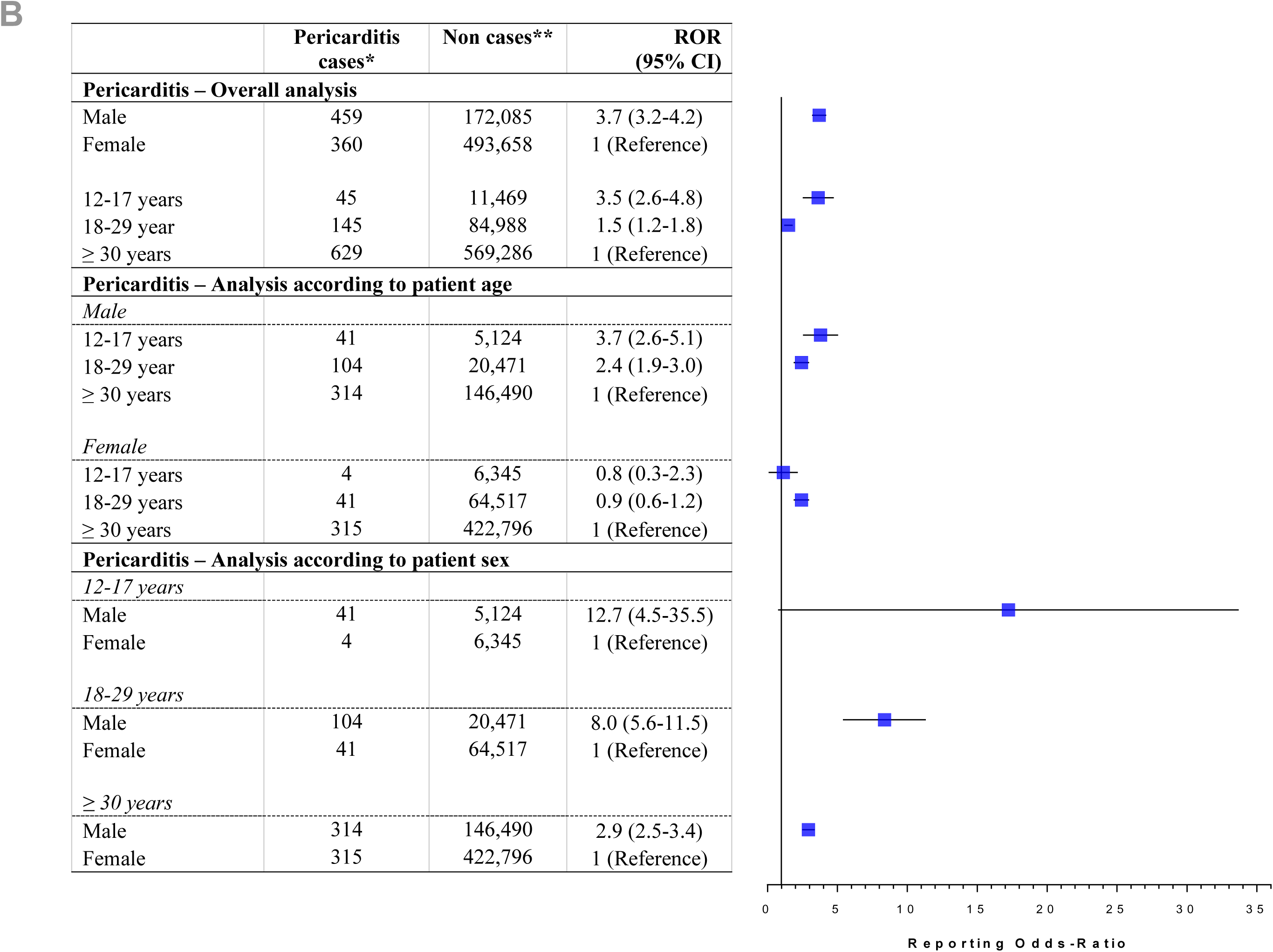
Comparison of reports of inflammatory cardiac disorders in COVID-19 mRNA vaccine recipients according sex and age within the WHO global safety database. ROR, Reporting odds-ratio; 95% CI, 95% confidence interval. Case–non-case approach is similar to case–control method but for the purposes of pharmacovigilance studies. Disproportionality in adverse drug reaction reporting between groups is expressed using reporting odds ratios (ROR) and their 95% confidence interval (95% CI). ROR is a ratio similar in concept to the odds ratio in case-control studies and corresponds to the exposure odds among reported cases of inflammatory cardiac disorders over the exposure odds among reported non-cases. An ROR over 1.0 suggests that inflammatory cardiac disorders are more frequently reported in COVID-19 mRNA vaccine recipients specific group compared with control group. We performed the main analysis and a secondary analysis stratified to age groups. * Inflammatory cardiac disorders cases were individual case safety reports containing the terms myocarditis or pericarditis as a reported preferred term according to the Medical Dictionary for Regulatory Activities (MedDRA, https://www.meddra.org/). Cases of myopericarditis have been considered as myocarditis. ** Non cases were reports containing any other reaction.

### Demographic and characteristics of cases from the U.S. VAERS

A total of 279 cases of inflammatory cardiac reaction with COVID-19 mRNA vaccines were reported in VAERS during the observed period. Of them, 157 (56%) myocarditis and 122 (44%) pericarditis cases were reported (**Table 2, Suppl. Table 1**). Patients were young belonging to the 18-39 age group (142, 51%), more commonly males (182, 65%). Adverse reactions occurred after the 1^st^ dose for 52 (19%), or after the 2^nd^ dose for 87 (31%) and unspecified for 140 (50%) of reported cases. COVID-19 disease prior to vaccination was reported for 18 (6%) patients. There was no concomitant COVID-19 reported. Although the median time to onset was similar for myocarditis and pericarditis, it tends to be longer for pericarditis (3 [2-4] days vs. 3 [2-9] days, respectively). Main symptoms are reported for myocarditis and pericarditis (respectively **Figure 3B and C**), with chest pain being most common. As illustrated in **Figure 5**, most cases of myocarditis experienced systemic reactogenicity signs (myalgia, fatigue, lymphedema on axillary, sweats and chills, fever, headache) appeared at day 1 (70, 45%), followed at day 3 to 4 by chest pain (107, 68%) and shortness of breath (26, 17%). Electrocardiogram (ECG) was abnormal for 68 (43%) patients with myocarditis and troponin was elevated for 41 (26%). The most common drugs reported to treat these myocarditis reactions included non-steroidal anti-inflammatory drugs (NSAID) (23, 15%) and colchicine (22, 14%). Corticosteroid therapy was infrequent (8, 5%). A total of 83 (53%) patients with reports of myocarditis were hospitalized, for a median duration of 2.5 days. **Figure 5** summarizes the typical patient management procedure for a myocarditis following mRNA COVID-19 vaccine.

**Figure 5.**
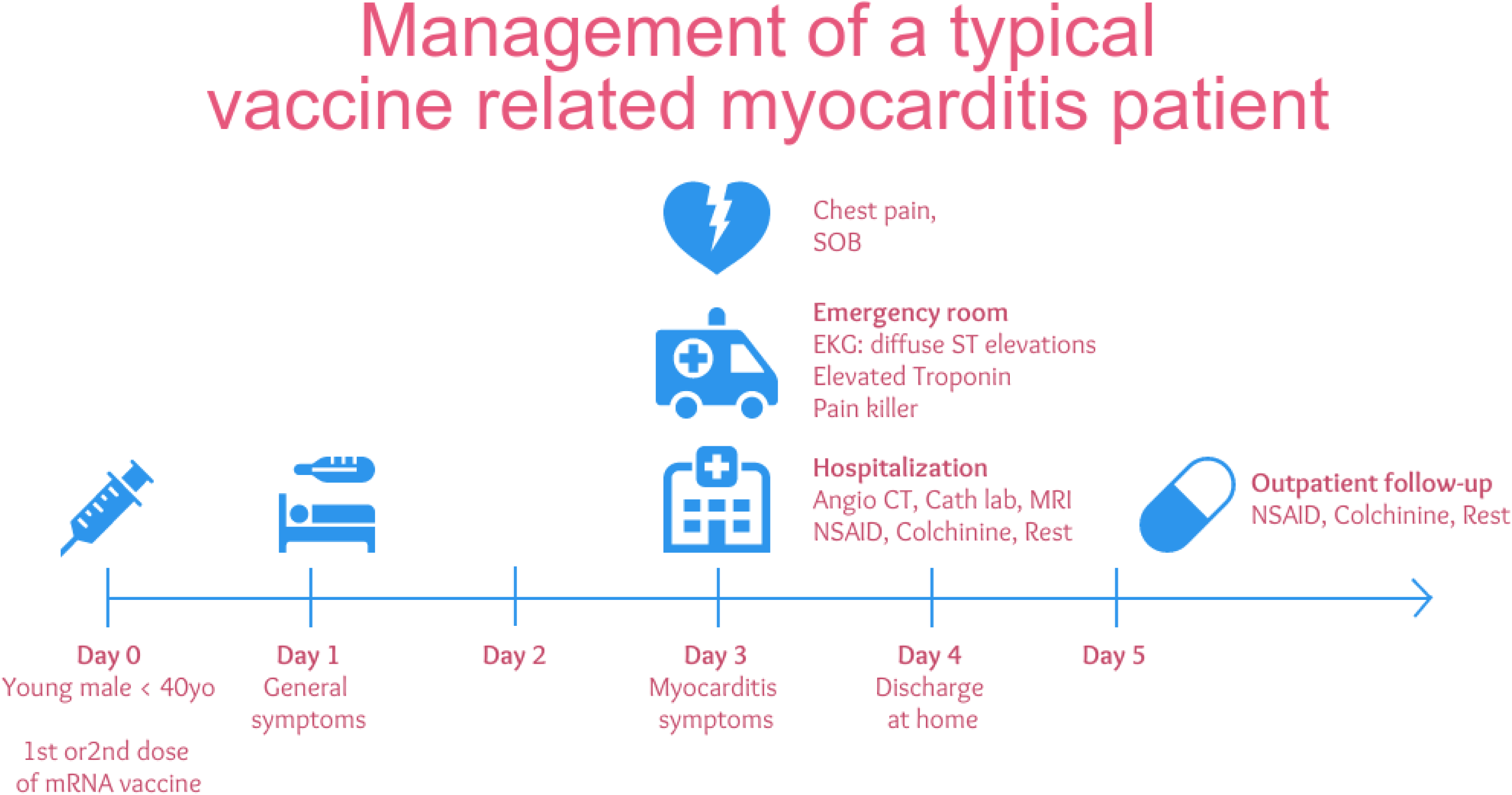
Management of a typical vaccine related myocarditis patient. The typical journey of patient experienced a myocarditis following an mRNACOVID-19 vaccine is described. Starting at day 0 after receiving 1^st^ or 2^nd^ dose of vaccine. Appearance of systemic reactogenicity symptoms at day 1, followed by chest pain and shortness of breath (SOB) at day 3, leading to consultation at the Emergency Room (ER), and hospitalization for a median of 2 days. SOB, shortness of breath; EKG, electrocardiogram; CT, computed tomography; NSAID, non-steroidal anti-inflammatory drug.

**Table 2.** Characteristics of acute inflammatory heart cases reported with COVID-19 mRNA vaccines within the VAERS classified by the type of reaction.

| **Reporting characteristics**<br>**(%) or median [IR]** | **Myocarditis**<br>**(n=157)** | **Pericarditis**<br>**(n=122)** | **Overall**<br>**(n=279)** |
| --- | --- | --- | --- |
| Vaccine type |  |  |  |
| - Tozinameran | 89 (57%) | 73 (60%) | 117 (42%) |
| - Elasmomeran | 68 (43%) | 49 (40%) | 162 (58%) |
| Sex |  |  |  |
| - Male | 114 (73%) | 68 (55%) | 182 (65%) |
| - Female | 38 (24%) | 52 (43%) | 90 (32%) |
| - Unknown | 5 (3%) | 2 (2%) | 7 (3%) |
| Age – ranges |  |  |  |
| - 12 - 17 years | 7 (4%) | 0 (0%) | 7 (3%) |
| - 18 - 29 years | 75 (48%) | 17 (14%) | 92 (33%) |
| - 30 – 39 years | 28 (18%) | 22 (18%) | 50 (18%) |
| - 40 - 49 years | 24 (15%) | 11 (9%) | 35 (13%) |
| - 50 - 59 years | 8 (5%) | 21 (17%) | 29 (10%) |
| - 60 - 64 years | 3 (2%) | 7 (6%) | 10 (4%) |
| - 65+ | 7 (4%) | 38 (31%) | 45 (16%) |
| - Unknown | 5 (3%) | 6 (5%) | 11 (4%) |
| COVID-19 prior to vaccination | 10 (6%) | 8 (7%) | 18 (6%) |
| After 1<sup>st</sup> dose | 22 (14%) | 29 (24%) | 52 (19%) |
| - Covid prior vaccination |  |  | 13 (25%) |
| - No Covid prior vaccination |  |  | 15 (29%) |
| - Unknown |  |  | 24 (46%) |
| After 2<sup>nd</sup> dose | 56 (36%) | 30 (25%) | 87 (31%) |
| - Covid prior vaccination |  |  | 2 (2%) |
| - No Covid prior vaccination |  |  | 34 (39%) |
| - Unknown |  |  | 51 (59%) |
| After unspecified dose | 79 (50%) | 63 (51%) | 140 (50%) |
| - Covid prior vaccination |  |  | 3 (2%) |
| - No Covid prior vaccination |  |  | 12 (9%) |
| - Unknown |  |  | 125 (89%) |
| Onset of the first symptoms – days | 3 [2-4] | 3 [2-9] | 3 [2-5] |
| Symptoms |  |  |  |
| - Systemic reactogenicity signs | 70 (45%) | 37 (30%) | 108 (39%) |
| - Chest Pain | 107 (68%) | 79 (65%) | 187 (67%) |
| - Shortness of breath | 25 (16%) | 31 (25%) | 57 (20%) |
| EKG |  |  |  |
| - Abnormal | 67 (43%) | 41 (34%) | 110 (39%) |
| - ST elevation | 41 (26%) | 13 (11%) | 54 (19%) |
| - Supraventricular arrhythmia | 3 (2%) | 13 (11%) | 16 (6%) |
| Elevated Troponin | 99 (63%) | 19 (16%) | 120 (43%) |
| Trans Thoracic Echocardiography |  |  |  |
| - Normal | 22 (14%) | 7 (6%) | 29 (10%) |
| - Decreased LVEF | 14 (9%) | 4 (3%) | 19 (7%) |
| CMR consistent with diagnosis | 29 (18%) | 1 (1%) | 30 (11%) |
| Complications |  |  |  |
| - CS or AHF | 9 (6%) | 1 (1%) | 10 (4%) |
| - Cardiac arrest | 2 (1%) | 0 (0%) | 2 (1%) |
| - Tamponade | 0 (0%) | 8 (7%) | 8 (3%) |
| Treatment |  |  |  |
| - NSAID | 23 (15%) | 0 (0%) | 43 (15%) |
| - Colchicine | 22 (14%) | 22 (18%) | 44 (16%) |
| - Corticosteroids | 8 (5%) | 3 (2%) | 12 (4%) |
| Hospitalization | 83 (53%) | 36 (30%) | 121 (43%) |
| Length of hospital stay – days | 2.5 [1-3] | 2 [1-4] | 2 [1-3] |

Data are presented as N (%) or median (IQR)
Data extracted from VAERS up to May 21<sup>st</sup> 2021.
AHF, Acute Heart Failure; CMR, Cardiac magnetic resonance ; CS, Cardiogenic Shock; EKG, electrocardiogram; LVEF, Left ventricular ejection fraction; NSAID, non-steroidal anti-inflammatory drug.

## Discussion

In this global pharmacovigilance retrospective observational study, we report here the largest case series to date on the clinical features of inflammatory heart reactions, myocarditis and pericarditis, following COVID-19 mRNA vaccine immunization. These cases mostly required hospitalization and some were life threatening, and occasionally fatal. Fatal cases occurred in about 1% of the patients, mostly older. Although our analysis cannot assess real incidence, these inflammatory heart reactions representing 31.8 per 10,000 of COVID-19 mRNA vaccine reports appeared to be rare. Regarding myocarditis, our global data show that they were highly likely to be reported in adolescents and young adults below 30 years old. Males has a high disproportionate reporting risk compared to females.

Overall clinical features from these databases are also consistent with the experience reported by the Israeli investigators. They have identified 110 myocarditis cases amongst their 5 million population who received 2 doses of the tozinameran vaccine.^14^ This translates to approximately 1 in 50,000 vaccine recipients, noting however the rates in young males were 5 to 25 times higher than expected. The investigators also observed a higher rate of incidence following the second dose, as well as 2 deaths in the cohort which may or may not be causally related. This overall pattern is consistent with features of traditional viral myocarditis, which occurs more commonly in younger male patients. Noteworthy, Israel’s spontaneous reports are not transmitted to VigiBase. Israel has been a leader in the current global mass immunization effort, due to the achievements of the campaign, a larger proportion of young people have been vaccinated compared to other countries using mRNA vaccines. The CDC reported that myocarditis reactions after vaccination with tozinameran vaccine were reported by adolescents aged 12–17 years to U.S. vaccine safety monitoring systems.^37^

Myocarditis and pericarditis have been previously reported after vaccinations.^8^ The most well studied example is that following smallpox vaccine, which in the relatively heathy U.S. military personnel had an incidence of myocarditis of 1 in 12,819 individuals.^38^ The complication rate was significantly decreased when the vaccine dose was subsequently reduced, suggesting a causal relationship, but it featured a live attenuated virus. Inflammatory cardiac reactions after inactivated virus vaccines such as influenza vaccines are also known to occur, but are generally considered as rare events, mainly occurring in young adults.^8^

Following the widespread adoption of community-based mitigation measures to reduce the transmission of SARS-CoV-2, the percentage of other respiratory diseases, such as influenza infection, has declined to historically low levels.^39, 40^ Therefore, we can hypothesize that the background risk for myocarditis is probably lower than usual. In this present study, distribution of myocarditis/pericarditis by sex and age was similar to previous cases described for other vaccines: mostly young adult males.^8^ Potential mechanisms for these observed reactions are currently unknown, but several hypotheses may be considered. The first is that incidence appears to be higher in situations related to higher immune reactivity (younger patient population, after second dose, etc.). This may be related to greater adaptive immune response in younger individuals, which in turn may lead to greater increases of CD4+ Th17+ cell populations, which predisposes individuals to developing myocarditis. It will be interesting to see if the recently reported microRNA diagnostic of Th17 activation in myocarditis is also positive in these patients.^41^ Second is the possibility that mRNA in these vaccines may enhance autoimmunity. ^42^ mRNA is known to be a self-adjuvant for innate immune responses, and this may help to explain their immunogenicity, and may also trigger excessive immune responses in some individuals, especially when there may be presence of a cross-reacting antigen. Optimization of mRNA dose delivered is needed to elicit appropriate immune response while minimizing undesirable inflammatory reactions, reassessing the dosing frequency and time interval between doses, as well as optimization of all ingredients associated with the messenger RNA, in order to achieve the most beneficial outcome of effective immunization with the least number of adverse effects to all different population demographics. No previous or concomitant COVID-19 disease before vaccination were reported in VAERS narratives. Some countries, such as France, recommend for individuals with previous SARS-CoV-2 infection to delay vaccination from three to six months after recovery, and to administer only one dose. Finally, with regards to COVID-19, it has been hypothesized that myocarditis can occur due to direct cell invasion via the spike protein interacting with the angiotensin-converting enzyme 2 (ACE2), which is widely expressed and prevalent in cardiomyocytes.^43, 44^ However, in case of COVID-19 related myocarditis, SARS-CoV-2 has not been found in cardiomyocytes, but only in the remaining myocardium, thus the cell injury was thought to be due to the generalized inflammatory response to COVID-19, part of which is Th1 activation.^45^ Hence in COVID-19 mRNA vaccine associated myocarditis, it appears unlikely that the translation of large amount of SARS-CoV-2 spike proteins through the vaccines may also engage the ACE2 and related receptors and play a role in reaction onset.

This retrospective pharmacovigilance analysis has some limitations and strengths. VigiBase and VAERS are based on spontaneous reports. Anyone can submit a report, including parents and patients, which are analyzed and documented by national or regional pharmacovigilance centers. Spontaneous reports will likely feature under-reporting of total real world cases and variable data quality, all of which are inherent to any pharmacovigilance system.^46^

For instance, previous prospective study of the incidence of myocarditis/pericarditis following smallpox and influenza vaccination suggested significant underestimation of true incidence of these complications with passive surveillance alone.^47^ Hence, one major limitation of spontaneous reporting is that it is not possible to assess incidence of inflammatory heart reactions following COVID-19 vaccination and total number of cases is highly underestimated. However, VigiBase, covering more than 90% of the world’s population, provides a unique opportunity to analyze rare adverse events at a global scale. Furthermore, although we cannot eliminate residual confounders, disproportionality analysis on VigiBase has proven its value in detecting safety signal or increased risk of events.^34^ Our analysis highlights an increased risk of myocarditis reporting in young males. On one hand, myocarditis has suffered from a notoriety bias in relation with media communication which has probably increase its reporting in May and June 2021 compared to other vaccine related events. On the other hand, because of an exceptionally high number of reports with these new COVID-19 mRNA vaccines that are associated to an important reactogenicity, non-cases reporting may be inflated as well, resulting in an underestimation of this risk. Finally, it is not likely that study groups in our disproportionality analysis, based on patient age and sex, may have been imbalanced in relation with these limitations. Sensitivity analyses showing consistent results strengthen our findings. Hence, data from VigiBase and VAERS reports should always be interpreted with caution, and consistency across datasets will be most important for validation of conclusions.^48^

We first report a global analysis on inflammatory cardiac reactions such as myocarditis/pericarditis occurring after COVID-19 mRNA vaccine. Compared to hundreds of millions of doses administered worldwide, these complications appeared to be rarely reported, although most of them required hospitalization. Reporting risk of myocarditis is highly increased in adolescents or young adults, and in males. Absolute risk may greatly increase with the achievements of the mass immunization campaign and the enlargement of vaccination to young adults. COVID-19 mRNA vaccine causality has been established by national authorities; further data will be needed to analyze this safety signal. Meanwhile, recommendations should be communicated to manage mass immunization in young adults and children, especially males. Some authors pledge for adapting the doses and number of doses needed.^49^ Benefits of COVID-19 vaccine still significantly outweigh their risks, including these rare inflammatory heart complications, given the current prevalence of SARS-CoV-2 infections in the world. As COVID-19 vaccine administration increases worldwide, clinicians and the public should be aware of the features of these inflammatory heart reactions, and follow appropriate monitoring and treatment procedures as with other cases of myocarditis.^50^ Maximal monitoring efforts by all relevant national and international agencies are needed to unlock key parameters to optimize future generations of vaccine platforms and dissemination to the general public resulting in maximum benefit potential. Guidelines are needed to take this specific risk into account and to adapt vaccination (dosage, timing and protocol) for younger people and males. In the future, as newer formulations are being developed, such as for vaccine boosters, the dose and timing for the at-risk cohort for cardiac complications should be considered to maximize the benefit to risk ratio.

## Data Availability

data available upon reasonable request

## Acknowledgments

VigiBase is a fully deidentified database maintained by the Uppsala Monitoring Center (UMC). The authors are indebted to the National Pharmacovigilance centres that contributed data. Information from VigiBase comes from a variety of sources, and the probability that the suspected adverse effect is drug-related is not the same in all cases. The information does not represent the opinion of the Uppsala Monitoring Center (UMC) or the World Health Organization and only reflects the authors’ opinion.

According to VigiBase access rules, no specific ethical approval is needed. VigiBase access is granted to national and regional pharmacovigilance centers such our teams.

## Sources of funding

This work is supported in part by research grants from the Canadian Institutes of Health Research, Heart & Stroke Foundation of Canada, Genome Canada, and University of Ottawa of Heart Institute.

## Disclosures

none

## Data availability

aggregated data of spontaneous reports are available at http://www.vigiaccess.org/.

## Author contributions

AB, LC and PL designed the study. AB and LC drafted the article and performed data extraction and statistical analysis. PL, MAK, TSK, GN, JR, MDD, MBVR, FS, JM and JMT critically reviewed the article. All the authors approved the final version of the article.

## ABBREVIATION LIST

CDC: Centers for Disease Control and Prevention
95%CI: Confidence Interval 95%
COVID-19: coronavirus disease
FDA: U.S. Food and Drug Administration
IQR: interquartile range
MedDRA: Medical Dictionary for Regulatory Activities
mRNA: messenger RNA
OR: Odds Ratio
ROR: Reporting Odds Ratio
VAERS: Vaccine Adverse Event Reporting System
WHO: World Health Organization

## FIGURE TITLES AND LEGENDS

**Supplemental Figure 1. Date of reporting and reaction of inflammatory heart reaction cases in VigiBase.**

## Notes

### Competing Interest Statement

The authors have declared no competing interest.

### Author Declarations

This study has obtained ethics approval from the Cochin university Hospital institutional review board (number AAA-2021-08039) in conformity with the French laws and regulations. All of the data used for analysis were de-identified, and only aggregate data are reported.

