## Supplemental Table 1 for "Cardiac Inflammation after COVID-19 mRNA Vaccines: A Global Pharmacovigilance Analysis"

**Suppl. Table 1. Characteristics of acute inflammatory heart cases reported with COVID-19 mRNA vaccines within the VAERS classified by the type of vaccine**

| **Reporting characteristics**  **(%) or median [IR]** | **Tozinameran vaccine**  **(n=162)** | **Elasomeran vaccine**  **(n=117)** | **Overall**  **(n=279)** |
| --- | --- | --- | --- |
| Sex   - Male - Female - Unknown | 102 (63%)  56 (35%)  4 (2%) | 80 (68%)  34 (29%)  3 (3%) | 182 (65%)  90 (32%)  7 (3%) |
| Age – ranges   - 12 - 17 years - 18 - 29 years - 30 – 39 years - 40 - 49 years - 50 - 59 years - 60 - 64 years - 65+ - Unknown | 7 (4%)  49 (30%)  27 (17%)  27 (17%)  21 (13%)  4 (2%)  23 (14%)  4 (2%) | 0 (0%)  43 (36%)  23 (20%)  8 (7%)  8 (7%)  6 (5%)  22 (19%)  7 (6%) | 7 (3%)  92 (33%)  50 (18%)  35 (13%)  29 (10%)  10 (4%)  45 (16%)  11 (4%) |
| Covid prior vaccination | 13 (8%) | 5 (4%) | 18 (6%) |
| Onset of the symptoms – days | 3 [2-6] | 3 [2-5] | 3 [2-5] |
| After 1^st^ dose   - Covid prior vaccination - No Covid prior vaccination - Unknown   After 2^nd^ dose   - Covid prior vaccination - No Covid prior vaccination - Unknown   After unspecified dose   - Covid prior vaccination - No Covid prior vaccination - Unknown | 32 (20%)  50 (31%)  80 (49%) | 20 (17%)  37 (32%)  60 (51%) | 52 (19%)  13 (25%)  15 (29%)  24 (46%)  87 (31%)  2 (2%)  34 (39%)  51 (59%)  140 (50%)  3 (2%)  12 (9%)  125 (89%) |
| Type of cardiac inflammatory injury   - Myocarditis - Myopericarditis - Pericarditis - Pleuropericarditis | 61 (38%)  28 (17%)  70 (43%)  3 (2%) | 48 (41%)  20 (17%)  46 (39%)  3 (3%) | 109 (39%)  48 (17%)  116 (42%)  6 (2%) |
| Symptoms   - General symptoms - Chest Pain - Shortness of breath | 67 (41%)  100 (62%)  31 (19%) | 41 (35%)  87 (74%)  26 (22%) | 108 (39%)  187 (67%)  57 (20%) |
| EKG   - Abnormal - ST elevation - Supraventricular arrhythmia | 62 (38%)  28 (17%)  8 (5%) | 48 (41%)  26 (22%)  8 (7%) | 110 (39%)  54 (19%)  16 (6%) |
| Elevated Troponin | 73 (45%) | 47 (40%) | 120 (43%) |
| Trans Thoracic Echocardiography   - Normal - Decreased LVEF | 16 (10%)  9 (6%) | 13 (11%)  10 (9%) | 29 (10%)  19 (7%) |
| CMR consistent with diagnosis | 15 (9%) | 15 (13%) | 30 (11%) |
| Complications   - CS or AHF - Cardiac arrest - Tamponade | 9 (6%)  1 (1%)  3 (2%) | 1 (1%)  1 (1%)  5 (4%) | 10 (4%)  2 (1%)  8 (3%) |
| Treatment   - NSAID - Colchicine - Corticosteroids | 29 (18%)  29 (18%)  7 (4%) | 14 (12%)  15 (13%)  5 (4%) | 43 (15%)  44 (16%)  12 (4%) |
| Hospitalization | 79 (49%) | 42 (36%) | 121 (43%) |
| Length of hospital stay – days | 2 [2-3] | 2 [1-3] | 2 [1-3] |

Data are presented as N (%) or median (IQR)

AHF, Acute Heart Failure; CMR, Cardiac magnetic resonance ; CS, Cardiogenic Shock; EKG, electrocardiogram; LVEF, Left ventricular ejection fraction*;* NSAID, non-steroidal anti-inflammatory drug.
