## Supplemental Table 2 for "Cardiac Inflammation after COVID-19 mRNA Vaccines: A Global Pharmacovigilance Analysis"

**Table S2. Reporting of myocarditis in COVID-19 mRNA vaccine recipients, and their reporting odds-ratios according to patient sex and age within the WHO global safety database: sensitivity analysis restricted to serious reports**

|  | **Myocarditis cases*** | **Non cases**** | **ROR (95% CI)** |
| --- | --- | --- | --- |
| **Myocarditis – Overall analysis** | | | |
| Male | 954 | 34,072 | 7.0 (6.1-8.0) |
| Female | 281 | 70,467 | 1 (Reference) |
| 12-17 years | 226 | 650 | 64.4 (54.1-76.7) |
| 18-29 year | 501 | 9,824 | 9.4 (8.3-10.7) |
| ≥ 30 years | 508 | 94,065 | 1 (Reference) |
| **Myocarditis – Analysis according to patient age** | | | |
| *Male* |  |  |  |
| 12-17 years | 205 | 354 | 55.8 (45.5-68.4) |
| 18-29 year | 423 | 2,329 | 17.5 (15.0-20.3) |
| ≥ 30 years | 326 | 31,389 | 1 (Reference) |
| *Female* |  |  |  |
| 12-17 years | 21 | 296 | 24.4 (15.3-38.9) |
| 18-29 years | 78 | 7,495 | 3.6 (2.7-47) |
| ≥ 30 years | 182 | 62,676 | 1 (Reference) |
| **Myocarditis – Analysis according to patient sex** | | | |
| *12-17 years* |  |  |  |
| Male | 205 | 354 | 8.2 (5.1-13.1) |
| Female | 21 | 296 | 1 (Reference) |
| *18-29 years* |  |  |  |
| Male | 423 | 2,329 | 17.5 (13.6-22.3) |
| Female | 78 | 7,495 | 1 (Reference) |
| *≥ 30 years* |  |  |  |
| Male | 326 | 31,389 | 3.6 (3.0-4.3) |
| Female | 182 | 6,2676 | 1 (Reference) |

Analysis restricted to serious reports (defined, according to the WHO, as the occurrence of death, life-threatening adverse event, inpatient hospitalisation or prolongation of an existing hospitalisation, significant disability or requirement of intervention to prevent any of these).

ROR, Reporting odds-ratio; 95% CI, 95% confidence interval.

* Myocarditis were individual case safety reports containing the terms myocarditis as a reported preferred term according to the Medical Dictionary for Regulatory Activities (MedDRA, <https://www.meddra.org/>). Cases of myopericarditis have been considered as myocarditis.

** Non cases were reports containing any other reaction.
