## Supplemental Table 3 for "Cardiac Inflammation after COVID-19 mRNA Vaccines: A Global Pharmacovigilance Analysis"

**Table S3. Reporting of myocarditis in COVID-19 mRNA vaccine recipients, and their reporting odds-ratios according to patient sex and age within the WHO global safety database: sensitivity analysis restricted to reports originating from healthcare professionals**

|  | **Myocarditis cases*** | **Non cases**** | **ROR (95% CI)** |
| --- | --- | --- | --- |
| **Myocarditis – Overall analysis** | | | |
| Male | 179 | 41,592 | 6.1 (4.7-7.8) |
| Female | 92 | 129,489 | 1 (Reference) |
| 12-17 years | 7 | 198 | 28.7 (13.3-61.9) |
| 18-29 year | 79 | 20,594 | 3.1 (2.4-4.1) |
| ≥ 30 years | 185 | 150,289 | 1 (Reference) |
| **Myocarditis – Analysis according to patient age** | | | |
| *Male* |  |  |  |
| 12-17 years | 7 | 85 | 28.0 (12.6-61.8) |
| 18-29 year | 63 | 4,512 | 4.7 (3.5-6.5) |
| ≥ 30 years | 109 | 36,995 | 1 (Reference) |
| *Female* |  |  |  |
| 12-17 years | - | 113 | - |
| 18-29 years | 16 | 16,082 | 1.5 (0.9-2.5) |
| ≥ 30 years | 76 | 113,294 | 1 (Reference) |
| **Myocarditis – Analysis according to patient sex** | | | |
| *12-17 years* |  |  |  |
| Male | 7 | 85 | - |
| Female | - | 113 | 1 (Reference) |
| *18-29 years* |  |  |  |
| Male | 63 | 4,512 | 14.0 (8.1-24.3) |
| Female | 16 | 16,082 | 1 (Reference) |
| *≥ 30 years* |  |  |  |
| Male | 109 | 36,995 | 4.4 (3.3-5.9) |
| Female | 76 | 113,294 | 1 (Reference) |

Analysis restricted to reports originating from healthcare professionals (i.e. physicians, pharmacists and other healthcare professionals).

ROR, Reporting odds-ratio; 95% CI, 95% confidence interval.

* Myocarditis were individual case safety reports containing the terms myocarditis as a reported preferred term according to the Medical Dictionary for Regulatory Activities (MedDRA, <https://www.meddra.org/>).

** Non cases were reports containing any other reaction.
