## Supplemental Table 4 for "Cardiac Inflammation after COVID-19 mRNA Vaccines: A Global Pharmacovigilance Analysis"

**Table S4. Reporting of pericarditis in COVID-19 mRNA vaccine recipients, and their reporting odds-ratios according to patient sex and age within the WHO global safety database: sensitivity analysis restricted to serious reports**

|  | **Pericarditis cases*** | **Non cases**** | **ROR (95% CI)** |
| --- | --- | --- | --- |
| **Pericarditis – Overall analysis** | | | |
| Male | 333 | 34,693 | 2.5 (2.2-3.0) |
| Female | 267 | 70,481 | 1 (Reference) |
| 12-17 years | 26 | 850 | 6.0 (4.0-8.9) |
| 18-29 year | 94 | 10,231 | 1.8 (1.4-2.2) |
| ≥ 30 years | 480 | 94,093 | 1 (Reference) |
| **Pericarditis – Analysis according to patient age** | | | |
| *Male* |  |  |  |
| 12-17 years | 24 | 535 | 5.8 (3.7-8.9) |
| 18-29 year | 66 | 2,686 | 3.1 (2.4-4.1) |
| ≥ 30 years | 243 | 31,472 | 1 (Reference) |
| *Female* |  |  |  |
| 12-17 years | 2 | 315 | 1.6 (0.4-6.7) |
| 18-29 years | 28 | 7,545 | 0.9 (0.6-1.4) |
| ≥ 30 years | 237 | 62,621 | 1 (Reference) |
| **Pericarditis – Analysis according to patient sex** | | | |
| *12-17 years* |  |  |  |
| Male | 24 | 535 | 7.1 (1.7-30.1) |
| Female | 2 | 315 | 1 (Reference) |
| *18-29 years* |  |  |  |
| Male | 66 | 2,686 | 6.6 (4.2-10.3) |
| Female | 28 | 7,545 | 1 (Reference) |
| *≥ 30 years* |  |  |  |
| Male | 243 | 31,472 | 2.0 (1.7-2.4) |
| Female | 237 | 62,621 | 1 (Reference) |

Analysis restricted to serious reports (defined, according to the WHO, as the occurrence of death, life-threatening adverse event, inpatient hospitalisation or prolongation of an existing hospitalisation, significant disability or requirement of intervention to prevent any of these).

ROR, Reporting odds-ratio; 95% CI, 95% confidence interval.

* Pericarditis were individual case safety reports containing the terms pericarditis as a reported preferred term according to the Medical Dictionary for Regulatory Activities (MedDRA, <https://www.meddra.org/>). Cases of myopericarditis have been considered as myocarditis.

** Non cases were reports containing any other reaction.
