## Supplemental Table 5 for "Cardiac Inflammation after COVID-19 mRNA Vaccines: A Global Pharmacovigilance Analysis"

**Table S5. Reporting of pericarditis in COVID-19 mRNA vaccine recipients, and their reporting odds-ratios according to patient sex and age within the WHO global safety database: sensitivity analysis restricted to reports originating from healthcare professionals**

|  | **Pericarditis cases*** | **Non cases**** | **ROR (95% CI)** |
| --- | --- | --- | --- |
| **Pericarditis – Overall analysis** | | | |
| Male | 162 | 41,609 | 4.1 (3.2-5.1) |
| Female | 124 | 129,457 | 1 (Reference) |
| 12-17 years | - | 205 | - |
| 18-29 year | 21 | 20,652 | 0.6 (0.4-0.9) |
| ≥ 30 years | 265 | 150,209 | 1 (Reference) |
| **Pericarditis – Analysis according to patient age** | | | |
| *Male* |  |  |  |
| 12-17 years | 0 | 92 | - |
| 18-29 year | 13 | 4,562 | 0.7 (0.4-1.2) |
| ≥ 30 years | 149 | 36,955 | 1 (Reference) |
| *Female* |  |  |  |
| 12-17 years | 0 | 113 | - |
| 18-29 years | 8 | 16,090 | 0.5 (0.2-1.0) |
| ≥ 30 years | 116 | 113,254 | 1 (Reference) |
| **Pericarditis – Analysis according to patient sex** | | | |
| *12-17 years* |  |  |  |
| Male | 0 | 92 | - |
| Female | 0 | 113 | 1 (Reference) |
| *18-29 years* |  |  |  |
| Male | 13 | 4,562 | 5.7 (2.4-13.8) |
| Female | 8 | 16,090 | 1 (Reference) |
| *≥ 30 years* |  |  |  |
| Male | 149 | 36,955 | 3.9 (3.1-5.0) |
| Female | 116 | 113,254 | 1 (Reference) |

Analysis restricted to reports originating from healthcare professionals (i.e. physicians, pharmacists and other healthcare professionals).

ROR, Reporting odds-ratio; 95% CI, 95% confidence interval.

* Pericarditis were individual case safety reports containing the terms pericarditis as a reported preferred term according to the Medical Dictionary for Regulatory Activities (MedDRA, <https://www.meddra.org/>). Cases of myopericarditis have been considered as myocarditis.

** Non cases were reports containing any other reaction.
