## Supplementary figures and images for "Cardiac Inflammation after COVID-19 mRNA Vaccines: A Global Pharmacovigilance Analysis"

### Supplemental Figure 1

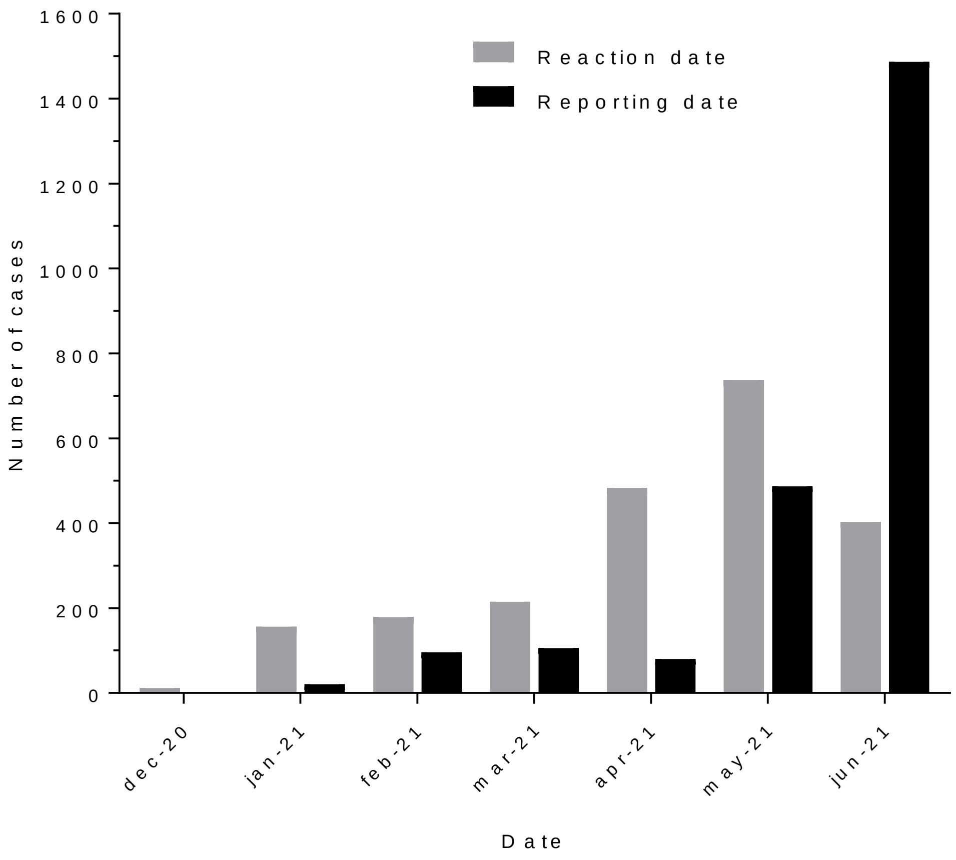
